# Trends in the Assessment, Treatment and Outcomes of Patients with Suspected Acute Coronary Syndrome

**DOI:** 10.64898/2026.07.03.26356908

**Authors:** Franz S Gruber, Alexander JF Thurston, Sara Hatam, Ryan Wereski, James Henderson, Iona Lyell, Yong Yong Tew, Deepak Harry, Samuel Chew, Zengyi Huang, Ziwen Li, Jennifer Daub, Jennifer Porteous, Alastair Hume, Arlene Casey, Dimitrios Doudesis, Nicholas L Mills, Atul Anand

**Affiliations:** DataLoch, NHS Lothian & University of Edinburgh, Edinburgh, UK; Centre for Population Health Sciences, University of Edinburgh, Edinburgh UK; BHF Centre of Research Excellence, Institute for Neuroscience and Cardiovascular Research, University of Edinburgh, UK

## Abstract

**Background:** Suspected acute coronary syndrome is a frequent Emergency Department (ED) presentation, requiring safe and efficient assessment. We interrogated long-term trends in whole population care for these patients using a new multi-centre regional registry.

**Methods:** The DataLoch Heart Disease Registry links relevant data from primary and secondary healthcare records, with national administrative data for patients registered within the Lothian Health Board region of Scotland (∼1M population). We included all adult patients presenting to secondary- or tertiary-care EDs in the region between 2014 and 2024, in whom high-sensitivity cardiac troponin was measured within 24 hours of presentation. Annual diagnostic rates for myocardial infarction, pharmacological and interventional management, and outcomes up to 1 year after ED presentation were studied. Logistic regression models were used to report change in annual trends for myocardial infarction, cardiac death, cardiovascular death and all-cause mortality, adjusted for age, sex, ethnicity, socioeconomic deprivation and comorbidity.

**Results:** Over 10 years, 117,142 consecutive patients (mean age 58 ± 18 years, 48% female, 6.6% with confirmed myocardial infarction) were included. Cardiac troponin testing increased year on year, from 61 per 1000 ED attendances in 2014 to 103 per 1000 in 2024 (*P*<0.001), but the proportion of patients admitted to hospital fell (59% in 2014 to 38% in 2024, *P*<0.001). Associated with these trends, the tested population had fewer cardiovascular risk factors and myocardial infarction incidence fell from 73 per 1000 tested patients in 2014 to 47 per 1000 in 2024 (adjusted odds ratio 0.62, 95% confidence intervals 0.56 to 0.69, *P*<0.001). In patients diagnosed with myocardial infarction, prescriptions of preventative therapies and numbers of revascularisation procedures were unchanged. After adjustment, no change over time was observed in one-year cardiac or cardiovascular mortality in those with a diagnosis of myocardial infarction.

**Conclusions:** ED testing using cardiac troponin has extended to a broader population at lower risk of myocardial infarction. Despite this trend, early rule-out pathways have reduced hospital admissions, without observable changes in outcomes for those with myocardial infarction.

**Clinical Perspective:** *What is new?:*

- Our registry approach captures all patients investigated for suspected acute coronary syndrome in Emergency Department settings for a regional population ∼1M people.
- Deep linkage across routine primary and secondary healthcare records combines patient characteristics, biomarker results, clinical diagnosis, interventional procedures, pharmacological treatment and adverse cardiovascular outcomes in 117,142 patients across more than a decade.

*What are the clinical implications?:*

- Cardiac troponin testing in the Emergency Department has been increasingly used in a wider population at lower risk of cardiac disease over the past decade.
- Despite this, the absolute number of patients diagnosed with myocardial infarction or undergoing revascularisation remained stable as did cardiovascular outcomes.
- Fewer patients required admission to hospital as accelerated early rule-out pathways were adopted, without evidence of harm.
- Data from this registry could support the design and evaluation of further refinements to diagnostic pathways for the assessment of patients with chest pain, help develop decision-support tools using machine learning, and inform wider public health policy.

## Introduction

Cardiovascular registries enable surveillance of quality of care and adherence to guidelines but can also establish whether findings from research studies in selected participants can be generalised to real world populations.^1^ A recent review identified 155 cardiac registries across 49 countries that inform and benchmark patient care.^2^ Large registries provide insights into conditions that are rare^3^ or difficult to study in a research setting,^4^ and have supported development of seminal clinical tools such as the Global Registry of Acute Coronary Events (GRACE) risk score.^5^

The diagnosis and management of myocardial infarction (MI) continues to evolve^6-9^ as our understanding and technologies improve.^10-14^ The past decade has seen the introduction of high sensitivity cardiac troponin (hs-cTn) assays, sex-specific diagnostic criteria, and rapid-rule out pathways.^15-18^ In addition, the recent SARS-CoV-2 pandemic changed the pattern of access to emergency departments for patients with suspected acute coronary syndrome (ACS).^19,20^

Current ACS registries are limited to confirmed cases of myocardial infarction (e.g. GRACE^21^, MINAP^22^, Euro Heart Survey^23^, ESC ACS^24^) or those under specialist cardiologist care (e.g. SWEDEHEART^25^). However, this represents only a fraction of the population who are assessed for signs and symptoms that could be related to ACS. This larger cohort forms a significant proportion of all Emergency Department (ED) attenders,^26^ but recent trends in care and clinical outcomes have not been systematically reported. Improved understanding could inform the design of new accelerated diagnostic pathways and help address wider system pressures that require new approaches to deliver safe but highly efficient care.^27^

We have developed the DataLoch Heart Disease registry to link healthcare data from multiple sources for a regional population in Scotland, relevant to a range of cardiovascular conditions. Using this regional registry, we identified consecutive attendances of patients with suspected ACS at EDs in secondary and tertiary care hospitals over the last decade. We present the trends in the assessment, management, and outcomes for patients investigated for suspected acute coronary syndrome.

### Material and Methods

Study design and participants

This retrospective, observational registry study included consecutive patients presenting with suspected acute coronary syndrome to an ED in a secondary or tertiary hospital in the Lothian region of South East Scotland (current population ∼1M) from 2014 to 2024. Patients were included if they were 18 years or older, and had hs-cTnI or hs-cTnT measured within 24 hours of ED attendance. Patients in whom ST-elevation myocardial infarction (STEMI) was diagnosed on a pre-hospital electrocardiogram and who were transferred directly to the cardiac catheterisation laboratory for emergent percutaneous coronary intervention without ED attendance were therefore not included. The registry was approved by the NHS Lothian delegated Caldicott Guardian.

## Data Linkage

The DataLoch Heart Disease Registry includes demographic data, health care observations, risk factors (e.g. smoking status, body mass index [BMI]) and clinical records extracted from participating primary care practice IT systems (Albasoft, Inverness, UK) and the Scottish Morbidity Record (SMR, Public Health Scotland) for hospital admissions. ED attendance data were sourced from the hospital electronic patient record that is used throughout the region (TrakCare, InterSystems Corporation, Boston, MA). Previous medical conditions and risk factors were defined from available primary or secondary care clinical records using Read or ICD10 codes listed in **Supplementary Table 1**. Relevant laboratory results were extracted from a combined database including tests requested in hospital and community settings (TrakCare, InterSystems Corporation, Boston, MA). Prescribing data was obtained from the Scottish national Prescribing Information System (PIS) that records regular and acute prescriptions dispensed in community pharmacies (all prescribed medications are provided without charge to residents in Scotland through the National Health Service; dispensed medications were identified with British National Formulary codes listed in **Supplementary Table 2**). Socioeconomic deprivation was determined using the Scottish Index of Multiple Deprivation (SIMD, Public Health Scotland) providing a geographical (post-code) measure of relative deprivation (https://www.simd.scot). Data related to deaths including cause were sourced from National Records of Scotland (NRS) statutory register of deaths. Participants who were not resident in Lothian at the the time of hospital attendance with suspected ACS were excluded from the registry.

### Measurement of cardiac troponin and thresholds for risk stratification

Cardiac troponin concentrations were measured in the course of clinical care by the hospital laboratories. From January 2014 until 27th October 2021, cardiac troponin was measured with the Abbott ARCHITECT-STAT hs-cTnI assay (Abbott Laboratories - limit of detection (LoD) of 1.2 ng/L; inter-assay coefficient of variation (CV) of <10% at 4.7 ng/L; 99th centile upper reference limit (URL) of 34 ng/L in men and 16 ng/L in women). After 27th October 2021 NHS Lothian hospital laboratories switched to the Roche Elecsys hs-cTnT assay (Roche Diagnostics - LoD 3 ng/L; CV <10%: 13 ng/L; male 99th centile URL: 16 ng/L, female 99th centile URL 9 ng/L). For each patient the maximum value of any hs-cTn test collected within 24h from ED presentation was taken, and categorised as ‘low’ if it was <5 ng/L, ‘intermediate’ if it was between 5 ng/L and the manufacturer’s sex-specific 99^th^ centile inclusive, and ‘high’ if it was greater than the manufacturer’s sex-specific 99^th^ centile value. Presenting symptoms (chest pain, dsypnoea, palpitations and syncope) were extracted from order set questions mandated when clinicians request a cardiac troponin test.

### Hospital admission and diagnosis of myocardial infarction

Myocardial infarction was determined from International Classification of Diseases (ICD-10) codes within national administrative data (SMR 01), where each hospital admission is summarised with up to six hierarchical codes. Patients were classified as having a myocardial infarction if their maximum cardiac troponin concentration was above the sex-specific 99th centile upper reference limit and the admission had ICD-10 codes of I21 or I22 in either of the first two positions in SMR 01. ST-segment elevation (STEMI) and non-ST-segment elevation myocardial infarction (NSTEMI) were classified using the 4th digit of I21/I22 codes. Length of stay and treating specialty was also determined from SMR 01 records. A full list of ICD-10 codes used to define the clinical diagnosis is provided in **Supplementary Table 1**.

### Mortality outcomes

Cardiac death was defined using ICD-10 diagnostic codes I05-I09, I20-I25, and I30-I52, limited to position 1 or 2 on the NRS death record. Cardiovascular death was defined using ICD-10 diagnostic codes I00-I99, also limited to position 1 or 2 on the death record.

### Revascularisation procedures

Interventions and procedures were extracted from OPCS version 4 codes within the hospital admission record; codes used to define revascularisation procedures can be found in **Supplementary Table 3**.

### Statistical analysis

Categorical variables are summarised as counts and percentages, and continuous variables as mean (SD) or median [IQR]. Categorical outcomes were compared with Pearson’s Chi-squared test. For analyses of outcomes and baseline characteristics, patients were counted once at their first presentation. Where data are tabulated by year, patients were included at their first presentation in the relevant calendar year. Trends in troponin testing were assessed using Mann-Kendall test. To assess comparative outcomes over time, logistic regression models were fitted for year-incidence of myocardial infarction, cardiac death, cardiovascular death or all-cause death, in each case using 2014 as the referent group. Odd ratios (OR) are reported with 95% confidence intervals (CI) and models were adjusted for age, sex, ethnicity, SIMD, previous myocardial infarction, previous stroke, diabetes, hypertension, chronic obstructive pulmonary disease (COPD) and asthma. Significance was considered at *P*<0.05. All statistical analysis was performed in R (version number 4.5, R Foundation for Statistical Computing).

## Results

### Cohort characteristics

From 2014 to 2024 there were 117,142 patients (48% female, mean age 58 ± 18 years) who presented with a suspected acute coronary syndrome, across a total of 195,817 attendances. Follow up to 30 days was available for all patients and for 104,468/117,142 (89%) to one year. Baseline characteristics at first presentation are shown in **Table 1**. Patients diagnosed with myocardial infarction at their first presentation (7,685 [6.6%]) were less likely to be female (2,634 [34%] *vs* 53,589 [49%], *P*<0.001) or to have normal kidney function (eGFR >60: 5,592 [73%] *vs* 92,125 [85%], *P*<0.001), and more frequently had cardiac risk factors (hypertension: 3,353 [44%] *vs* 33,040 [30%], *P*<0.001; current/previous cigarette smoking: 5,192 [68%] *vs* 64,314[58%], *P*<0.001). Cardiac risk factors in the population tested for suspected ACS decreased in prevalence over the course of the study period (**Supplementary Table 4**).

**Table 1:**
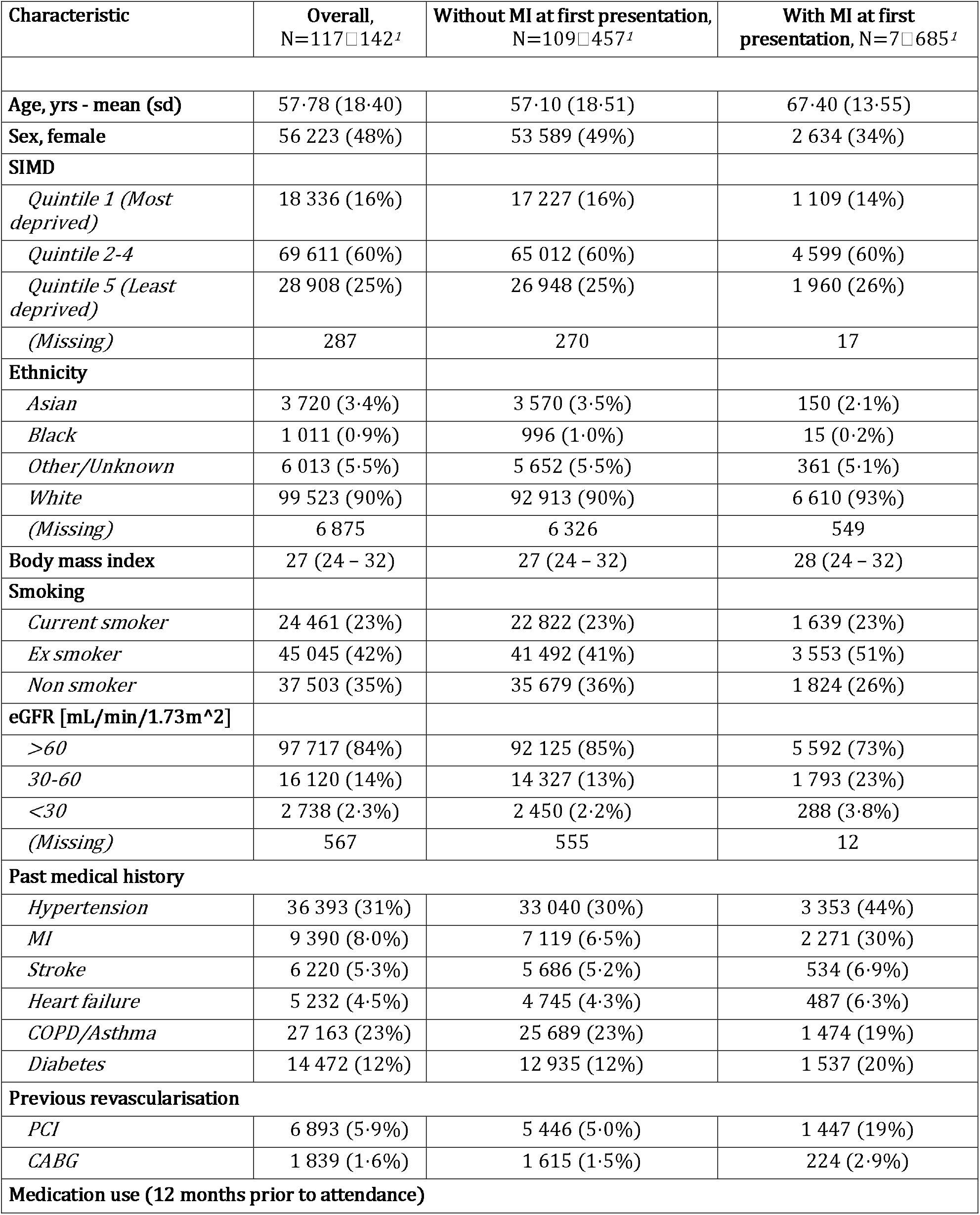
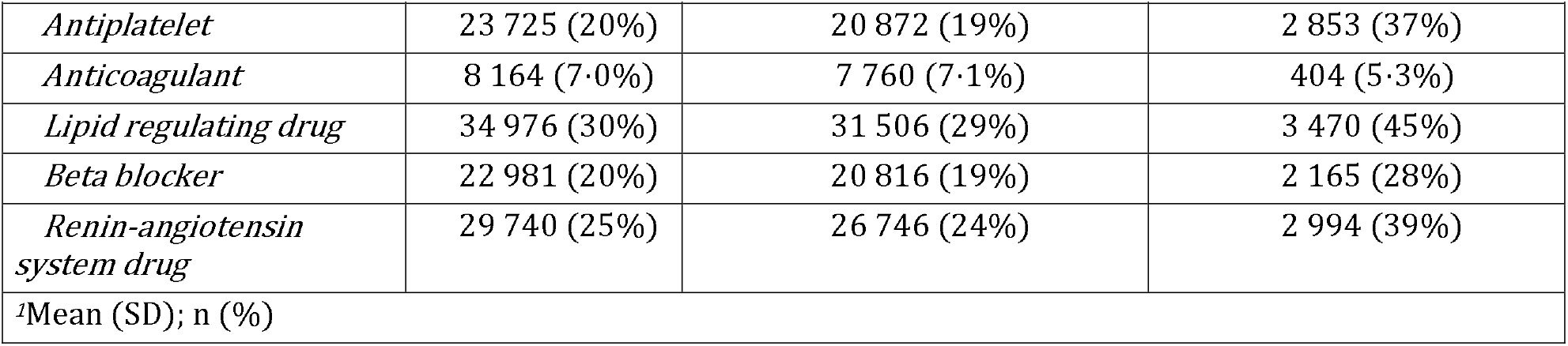
Baseline characteristics at first presentation, stratified according to diagnosis of MI.

### Trends in cardiac troponin testing

From 2014 to 2021 cardiac troponin testing increased from 481 to 825 patients tested per 10,000 total ED attendances (**Figure 1A-B**, *P*<0.001). On average, chest pain was the most commonly recorded symptom in ∼85% of attendances (**Supplementary Figure 1**). In the early phase of the COVID-19 pandemic, the overall rate of ED attendances dropped twice (April 2020 and February 2021) coinciding with nationwide ‘lockdowns’28. While the total number of attendances with troponin testing fell in April 2020, these cases represented a greater proportion of all ED patients. Following the switch from a hs-cTnI to hs-cTnT assay (October 2021), monthly attendances with cardiac troponin testing remained static (Mann-Kendall test for trend, *P*=0.23). However, fewer patients had low cardiac troponin concentrations below local clinical early rule-out threshold (<5 ng/L), and more were identified with myocardial injury (above the sex-specific 99th centile, **Figure 1C**). Despite this, annual incidence of myocardial infarction decreased after this change, continuing a decreasing trend over time from 73 per 1,000 suspected ACS attendances in 2014 to 47 per 1000 attendances in 2024 (adjusted OR 0.62, 95% CI 0.56-0.69, *P*<0.001).

**Figure 1.**
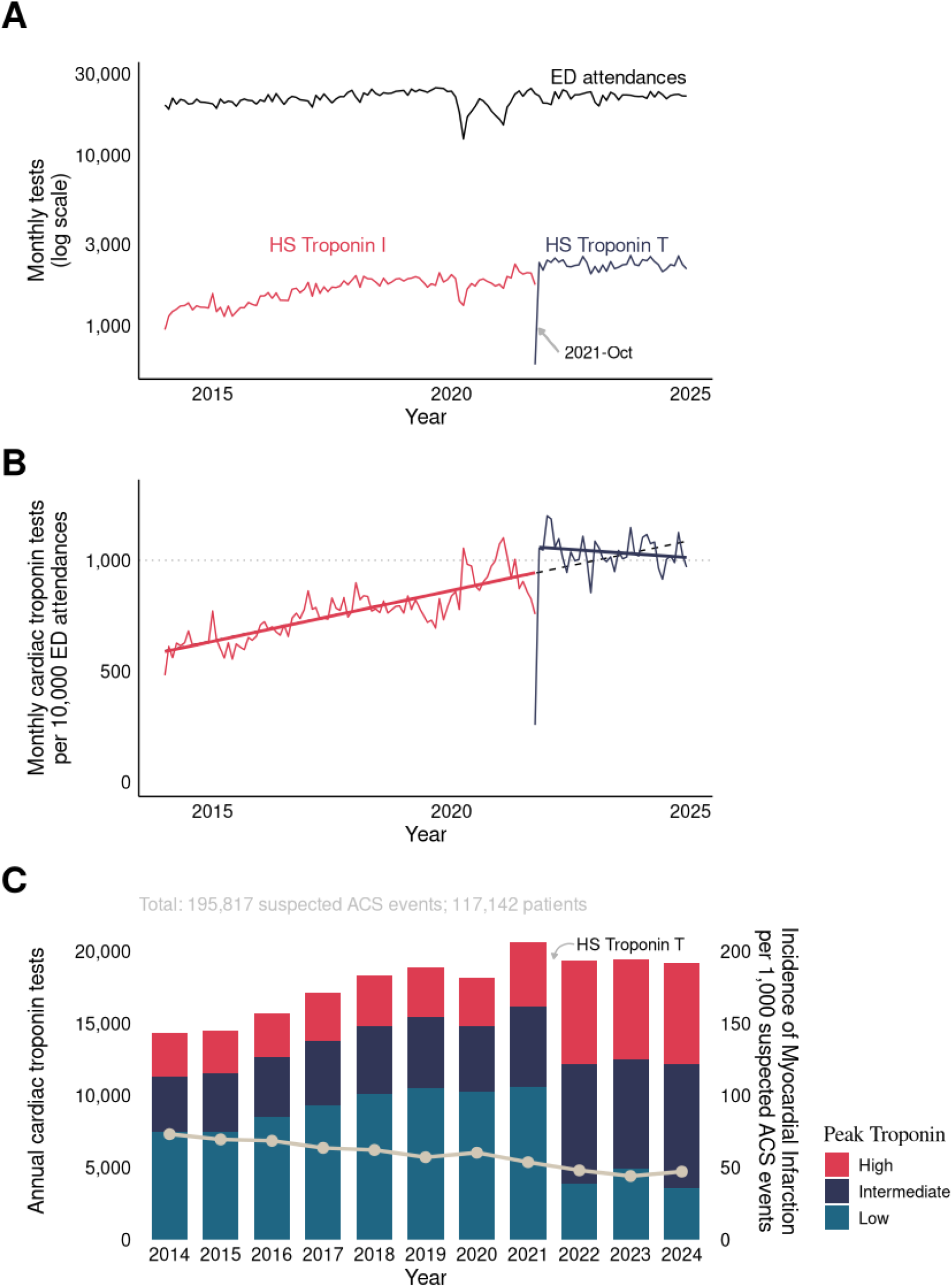
(A) Patients tested monthly for cardiac troponin in Lothian Emergency Departments, alongside monthly Emergency Department attendances (any attendance reason). Colors: HS Troponin I (red), HS Troponin T (blue), ED attendances (black). (B) Patients tested for cardiac troponin per 10,000 ED attendances. Lines: reference line (black dotted), linear fit through the whole time period (black dashed), linear fits through HS Troponin I period (red solid), linear fit through HS Troponin T period (blue solid). First day of HS Troponin T testing was omitted from the linear fit. (C) Absolute numbers of ED Attendances with at least one cardiac troponin test, segmented to show the proportions <5ng/L (low), 5-99th centile (intermediate), >99th sex-specific centile (high) (based on maximal troponin value within 25 hours since presentation). Grey line shows the incident rate of myocardial infarction (per 1,000 tested patients) arising from these episodes.

### In hospital management

Of the 7,685 patients receiving a diagnosis of myocardial infarction at their first presentation, 5,546 [72%] underwent angiography, with 4,227 [55%] receiving percutaneous coronary intervention (PCI) within 30 days from attendance (**Table 2**). A further 285 [3·8%] coronary artery bypass grafting (CABG) procedures were performed in this timeframe. Patients were most often managed by specialist cardiology services, and median length of stay remained stable at 3 days throughout the study period (**Figure 2A**). Annual rates of coronary angiography, intervention and cardiac medication prescriptions remained consistent across the study period (**Figure 2B-C**). Patients that were managed under cardiology services were more likely to receive dual antiplatelet therapy, lipid lowering drugs, beta blockers or renin-angiotensin-aldosterone (RAAS) inhibitors within 1 year of diagnosis (all *P* <0.001 compared to non-specialist management).

**Table 2:** Cardiovascular outcomes following first presentation with suspected acute coronary syndrome.

| Characteristic | Overall,<br>N=117□142<sup>1</sup> | Without MI at first presentation,<br>N=109□457<sup>1</sup> | With MI at first presentation,<br>N=7□685<sup>1</sup> |
| --- | --- | --- | --- |
| *Admitted* | 50 282 (43%) | 42 597 (39%) | 7 685 (100%) |
| *ED (hours)* | 4 (3 – 5) | 4 (3 – 5) | 4 (3 – 5) |
| *ED + inpatient (days)* | 0 (0 – 2) | 0 (0 – 1) | 3 (2 – 5) |
| *Angiography* | 7 932 (6.8%) | 2 386 (2.2%) | 5 546 (72%) |
| **Revascularisation within 30 days from attendance** |  |  |  |
| *PCI* | 5 247 (4.5%) | 1 020 (0.9%) | 4 227 (55%) |
| *CABG* | 472 (0.4%) | 187 (0.2%) | 285 (3.7%) |
| **Any reattendance from discharge** |  |  |  |
| *30 days* | 14 379 (12%) | 13 052 (12%) | 1 327 (17%) |
| *1 year* | 46 118 (39%) | 42 797 (39%) | 3 321 (43%) |
| **Reattendance with suspected ACS from discharge** |  |  |  |
| *30 days* | 5 166 (4.4%) | 4 254 (3.9%) | 912 (12%) |
| *1 year* | 15 117 (13%) | 13 068 (12%) | 2 049 (27%) |
| **Reattendance with MI from discharge** |  |  |  |
| *30 days* | 334 (0.3%) | 203 (0.2%) | 131 (1.7%) |
| *1 year* | 982 (0.8%) | 667 (0.6%) | 315 (4.1%) |
| **Death (all-cause) from attendance** |  |  |  |
| *inpatient* | 2 319 (2.0%) | 1 820 (1.7%) | 499 (6.5%) |
| *30 days* | 2 975 (2.5%) | 2 354 (2.2%) | 621 (8.1%) |
| *1 year* | 6 226 (5.3%) | 5 284 (4.8%) | 942 (12%) |
| **Death (cardiovascular) from attendance** |  |  |  |
| *inpatient* | 1 515 (1.3%) | 1 068 (1.0%) | 447 (5.8%) |
| *30 days* | 1 907 (1.6%) | 1 359 (1.2%) | 548 (7.1%) |
| *1 year* | 3 585 (3.1%) | 2 832 (2.6%) | 753 (9.8%) |
| **Death (cardiac) from attendance** |  |  |  |
| *inpatient* | 1 151 (1.0%) | 720 (0.7%) | 431 (5.6%) |
| *30 days* | 1 462 (1.2%) | 936 (0.9%) | 526 (6.8%) |
| *1 year* | 2 657 (2.3%) | 1 967 (1.8%) | 690 (9.0%) |
| <sup>1</sup>n (%); Median (IQR) |  |  |  |

**Figure 2.**
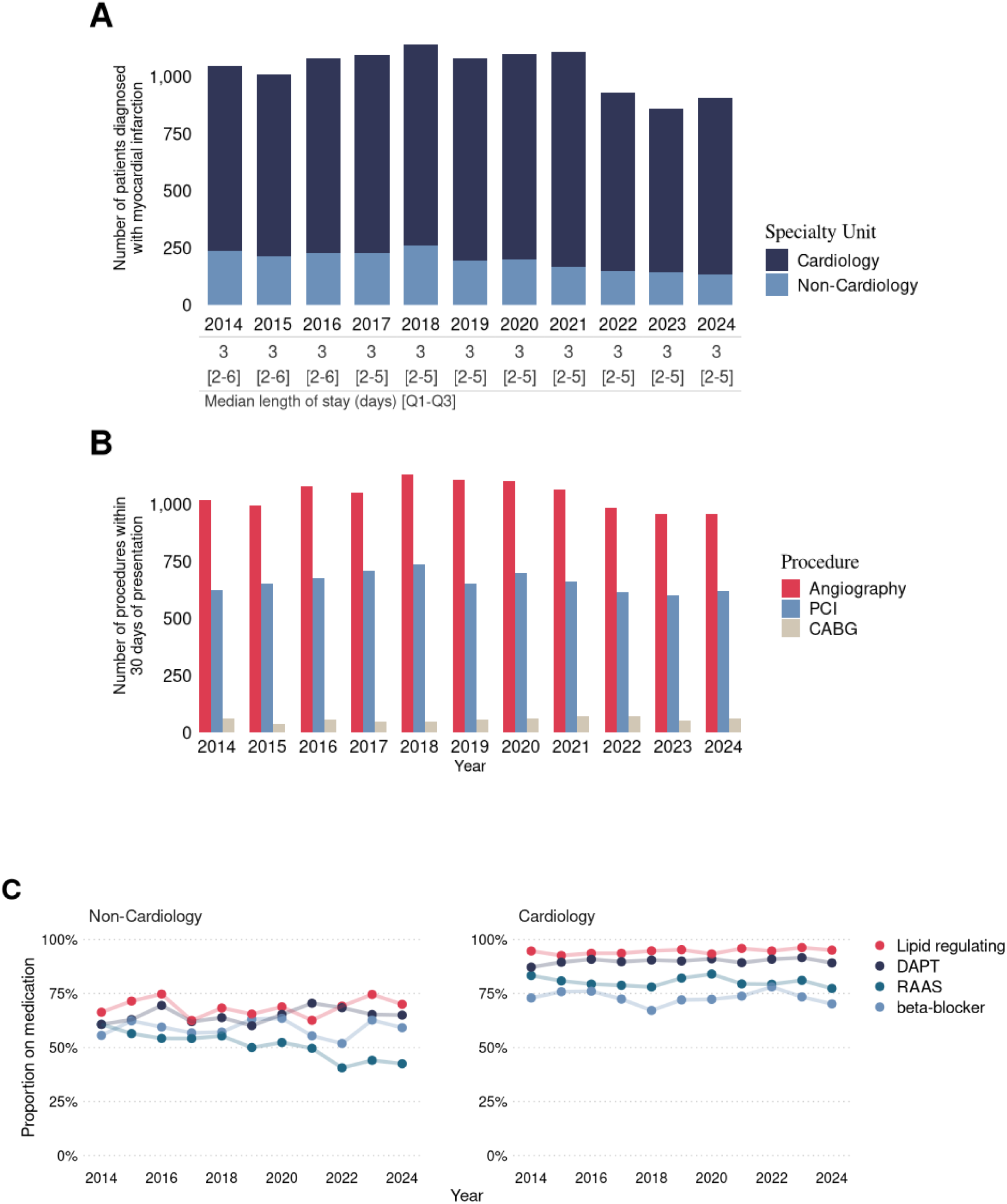
(A) Absolute number of cases diagnosed with myocardial infarction per year, split by management under a cardiology specialty code *vs* non-cardiology. (B): Absolute number of coronary angiograms performed per year among registry participants. (C) Annual proportion of patients receiving dual anti-platelet or anticoagulant therapy (DAPT-dark blue), renin-angiotensin-aldosterone signalling inhibition (RAASi-teal), lipid lowering therapy (red), or beta-blockers (light blue) per year in the 12 months following myocardial infarction.

Of patients without a diagnosis of myocardial infarction, 42,597 [39%] were still admitted to hospital, but this proportion fell from 56% in 2014 to 34% in 2024. The median length of stay for these patients was shorter (0 (0 – 1) days), and few of these patients underwent angiography (2,386 [2·2%]) or revascularization (PCI: 1,020 [0·9%], CABG: 187 [0·2%]) within 30 days of presentation (**Table 2**). Length of stay in the ED rose in the years after the COVID-19 pandemic for patients both with and without a diagnosis of myocardial infarction (**Supplementary Tables 5 & 6**),

### Modeling trends in diagnosis and outcomes

The odds of receiving a diagnosis of myocardial infarction decreased over time (adjusted OR 0.64 in 2024, 95% CI 0.57-0.72, using 2014 as reference, **Figure 3A**). Patients diagnosed with myocardial infarction were more likely to reattend subsequently with a suspected acute coronary syndrome than those without myocardial infarction (912 [12%] vs. 4,254 [3·9%] within 30 days; 2,049 [27%] *vs* 13,068 [12%] within 1 year from index presentation, **Table 2**). A total of 942 (12%) patients died within one year of a myocardial infarction (690 [9·0%] from a cardiac cause). The risk of all-cause mortality at one year following presentation was also high in patients who were not diagnosed with myocardial infarction at their first presentation (5,284 [4·8%]). However, only 1,967 (1·8%]) died from cardiac causes. Cardiac-specific and all-cause mortality rates decreased over the study period for both those with and without a diagnosis myocardial infarction (**Figure 3, Supplemental Tables 5 & 6**). However, once adjusted for demographics and comorbidity, no reductions were observed in patients with myocardial infarction (cardiac death adjusted OR 0.88 in 2023, 95% CI 0.58-1.30, *P*=0.52, using 2014 as reference) or without (adjusted OR 0.88 in 2023, 95% CI 0.71-1.09, *P*=0.25).

**Figure 3.**
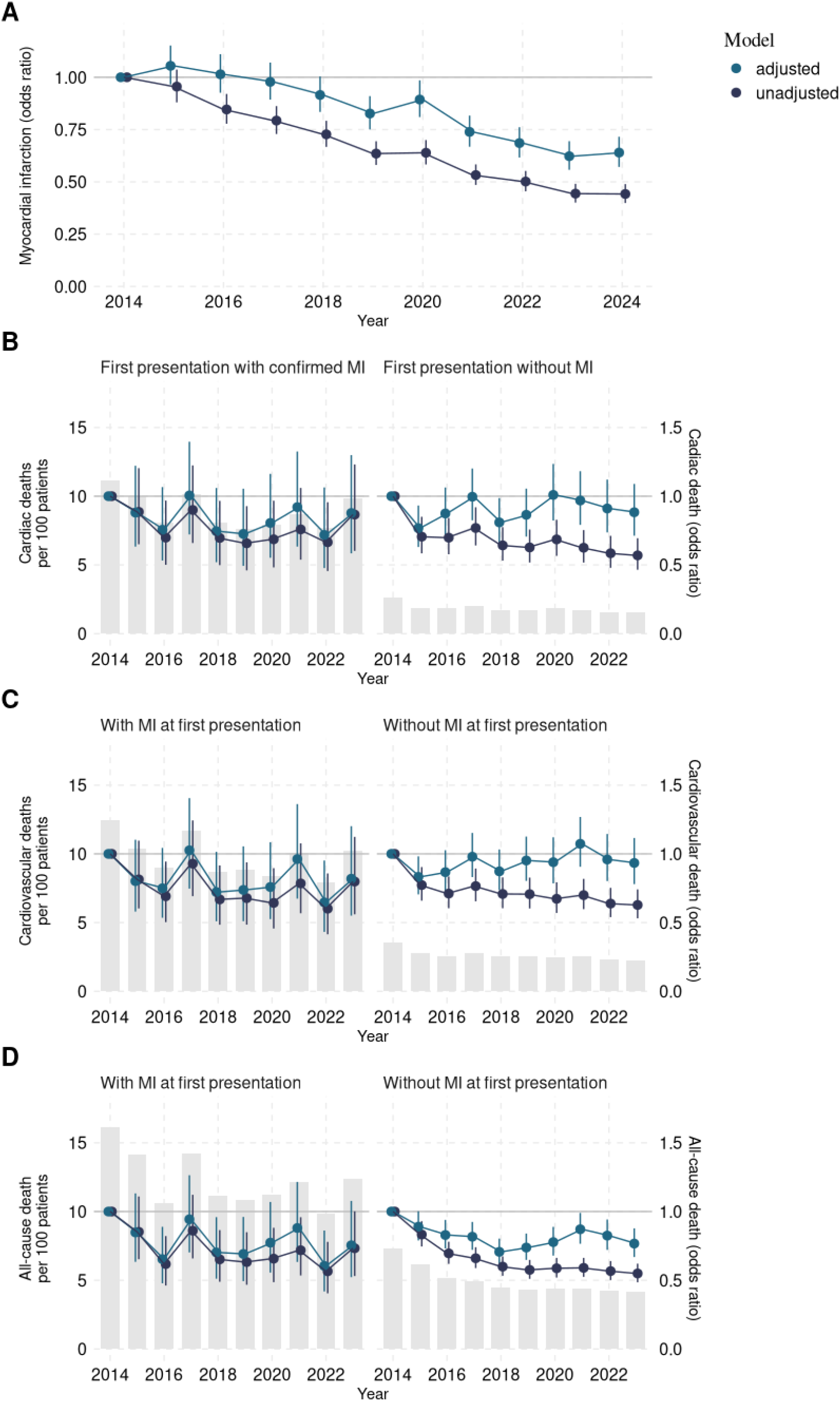
Trends in the diagnosis of myocardial infarction (panel A) and one year cardiac mortality (panel B), cardiovascular mortality (panel C) and all-cause mortality (panel D), stratified according diagnosis at first presentation with suspected ACS. Grey bars show the absolute number of cause-specific deaths at one year. Dark blue lines show the trend in odds ratio associated with each year, indexed to 2014, from an unadjusted logistic regression model. Teal lines show the change in odds ratio when the model is adjusted for age, sex and cardiovascular risk factors.

## Discussion

We have used robustly linked routine electronic healthcare record data to understand the characteristics, investigations, diagnoses, management and outcomes for a decade of clinical practice assessing patients with suspected acute coronary syndrome. We report important findings with implications for clinical practice, healthcare systems and the future refinement of accelerated diagnostic pathways for myocardial infarction. Cardiac troponin testing rates have doubled for ED attendances over this period, with spread to populations at lower objective cardiovascular risk and pre-test probablility of myocardial infarction. Despite this expansion, accelerated early rule-out pathways have lowered hospital admission in both relative and absolute terms over time. However, with greater testing of lower risk individuals the relative incidence of myocardial infarction in those tested has fallen, and invasive coronary procedure rates are unchanged. There has been no sustained improvement in cardiac or cardiovascular death despite falling all-cause mortality rates in the whole population tested over this period. Taken together these registry data reveal important system trends in ED management that reflect less-specific testing for ACS since the introduction of high-sensitivity cardiac troponin assays.

Our registry covers a period from the introduction of high-sensitivity cardiac troponin assays into clinical practice. This enabled an accelerated clinical decision pathway to be introduced in 2015^17^ with early rule-out for many patients using a single sample cardiac troponin testing strategy. Our data shows marked and sustained reduction in hospital admissions for those without myocardial infarction since this change, from the majority of patients attending ED with suspected ACS to just one third. Reattendance in this group was stable, and mortality rates decreased in line with the overall trend observed across the study period. Our registry shows the true long-term clinical benefit and safety of the application of accelerated rule-out pathways that has been made possible by greater precision at low cardiac troponin concentrations using high-sensitivity cardiac troponin assays over the last decade. However, these effective pathways are not immune to wider system pressures;^29^ despite the opportunity to discharge a significant proportion of patients with a single blood test, the ED length of stay for patients with and without myocardial infarction has increased in the last three years. These results may also reflect an assay switch to high-sensitivity cardiac troponin T, following which substantially fewer patients had presentation levels suitable for single test rule-out to exclude myocardial infarction under local guidelines. Troponin testing plateaued after this assay switch and diagnoses of myocardial infarction fell, perhaps as clinicians felt less confident that hs-cTnT concentrations above the upper reference limit were due to myocardial infarction^30^. In keeping with this, the number of STEMI diagnoses made in ED remained stable after this change, whereas diagnoses of NSTEMI fell. Risk of cardiac death did not rise in patients with myocardial infarction excluded, suggesting this assay switch did not increase missed cardiac events. Equally, no consistent trend in cardiac death was observed for those with myocardial infarction after this change.

Presentations with suspected acute coronary syndrome and diagnoses of myocardial infarction were relatively unaffected by the COVID-19 pandemic, and there was no clear excess mortality during this period, despite fears that patients would avoid seeking medical attention or strain on wider hospital services would impact quality of care^31^. Risk of mortality due to non-cardiac causes remained elevated in those patients who were not diagnosed with myocardial infarction, with one in twenty patients dead at one year. This highlights the need for clinical decision pathways for unselected patients with chest pain to emphasise the importance of seeking an alternate diagnosis when myocardial infarction has been excluded.

Our study reports a lower prevalence of myocardial infarction, use of angiography and revascularisation than observed in other major registries of patients with myocardial infarction or chest pain presentations.^32-34^ We believe this reflects the strength of our approach to include consecutive ED presentations, in contrast to other registries which are contingent on an admission under specialist cardiology services, confirmation of myocardial infarction or where inclusion of patients without myocardial infarction is optional. Our registry informs interpretation of findings in these studies by characterising an unselected population with suspected acute coronary syndrome, which may give a more accurate estimate of prevalence.

There are some limitations which should be considered. The diagnosis of myocardial infarction was based on ICD-10 coding, which does not allow stratification of patients according to the sub-type of myocardial infarction under the Fourth Universal Definition of Myocardial Infarction.^7^ Patients with type 2 myocardial infarction require different investigation and management strategies, and this may account for the differing rates of preventative therapy prescription in patients cared for by cardiologists and non-specialists. This group are known to have more comorbidities and poorer outcomes,^35,36^ and distinguishing these patients would have informed our analysis. Further research is needed to understand if lower rates of secondary preventative medications in patients cared for by non-specialists are fully explained by different management strategies for type 2 myocardial infarction, or if this represents a true treatment gap. In our region, patients with pre-hospital ST-segment elevation are typically conveyed directly to the cardiac catheterisation laboratory and do not attend the ED, so were not captured in our registry. The total prevalence of myocardial infarction is therefore higher in this group than we report in our cohort,^37^ and it is appropriate to evaluate the diagnostic journey of a STEMI population separately.

This Heart Disease Registry has the potential to be utilised as a platform supporting data-enabled trials with smart outcome capture in near real-time. In future, the registry will be augmented with adjudicated diagnoses from research trials,^15,17^ extension into chronic coronary syndromes and heart failure, and by linkage with electrocardiograms and cardiac imaging, alongside appropriate computing infrastructure to support deep learning. This will create a powerful resource to support the development, validation and evaluation of new diagnostic tools, care pathways and algorithms that provide clinical decision support for cardiac care. The registry can be accessed remotely via a secure data environment by approved researchers following successful application to DataLoch (https://www.dataloch.org).

In conclusion, the testing for acute coronary syndrome in our region has increased over the past decade, while diagnostic rates for myocardial infarction have fallen and clinical pathways have more efficiently discharged ED attenders. Our registry-based analysis represents a new and ongoing resource to inform our understanding of the assessment of suspected ACS, and to optimise systems of care to improve diagnosis and management for this important patient group.

## Data Sharing

The study used routine electronic health care data sources that linked, de-identified and aggregated individual patient level data that is held in a Trusted Research Environment by DataLoch (https://dataloch.org/). The study data and analysis code can be accessed by individuals who have undertaken the necessary governance training on application to DataLoch.

## Acknowledgements

This work uses data provided by patients and collected by the NHS as part of their care and support. This project has been facilitated by the DataLoch service. DataLoch enables access to de-identified extracts of health care data from the South-East Scotland region to approved applicants: dataloch.org.

## Sourcees of funding

This study was funded through a Programme Grant from the British Heart Foundation (RG/20/10/34966). FSG and AC are supported by a DARE UK grant (UKRI3005). AJFT is supported by a Clinical Research Training Fellowship (FS/CRTF/25/24735) from the British Heart Foundation. IL is supported by a Clinical Research Training Fellowship (FS/CRTF/25/24788) from the British Heart Foundation. ZL is supported by a Project Grant (PG/25/12298) from the British Heart Foundation. AC is funded by the Vivensa Foundation [PF2302\2]. NLM is supported by the British Heart Foundation through a Chair Award (CH/F/21/90010), a Programme Grant (RG/F/25/110169) and a Research Excellence Award (RE/24/130012).

