## Supplemental Material for "Trends in the Assessment, Treatment and Outcomes of Patients with Suspected Acute Coronary Syndrome"

Short Title: Trends in suspected Acute Coronary Syndrome

Franz S Gruber PhD^1,2^*, Alexander JF Thurston MD^3^*, Sara Hatam MSc^1^, Ryan Wereski MD^3^, James Henderson MD^3^, Iona Lyell MD^3^, Yong Yong Tew MD^3^, Deepak Harry MD^3^, Samuel Chew MD^3^, Zengyi Huang^1^, Ziwen Li MD PhD^3^, Jennifer Daub^1^, Jennifer Porteous^1^, Alastair Hume BSc (Hons)^1^, Arlene Casey PhD^1,2^, Dimitrios Doudesis PhD^3^, Nicholas L Mills MD PhD^1,3†^, Atul Anand MD PhD^1,3†^

1 DataLoch, NHS Lothian & University of Edinburgh, Edinburgh, UK
2 Centre for Population Health Sciences, University of Edinburgh, Edinburgh UK
3 BHF Centre of Research Excellence, Institute for Neuroscience and Cardiovascular Research, University of Edinburgh, UK

STROBE checklist

|  | Item No | Recommendation | Page |
| --- | --- | --- | --- |
| **Title and abstract** | 1 | (*a*) Indicate the study’s design with a commonly used term in the title or the abstract | 2 |
|  |  | (*b*) Provide in the abstract an informative and balanced summary of what was done and what was found | 2 |
| Introduction | | |  |
| Background/rationale | 2 | Explain the scientific background and rationale for the investigation being reported | 5-6 |
| Objectives | 3 | State specific objectives, including any prespecified hypotheses | 6 |
| Methods | | |  |
| Study design | 4 | Present key elements of study design early in the paper | 6-7 |
| Setting | 5 | Describe the setting, locations, and relevant dates, including periods of recruitment, exposure, follow-up, and data collection | 6-10 |
| Participants | 6 | (*a*) Give the eligibility criteria, and the sources and methods of selection of participants. Describe methods of follow-up | 6-10 |
|  |  | (*b*) For matched studies, give matching criteria and number of exposed and unexposed | NA |
| Variables | 7 | Clearly define all outcomes, exposures, predictors, potential confounders, and effect modifiers. Give diagnostic criteria, if applicable | 6-10 |
| Data sources/ measurement | 8* | For each variable of interest, give sources of data and details of methods of assessment (measurement). Describe comparability of assessment methods if there is more than one group | 6-10 |
| Bias | 9 | Describe any efforts to address potential sources of bias | 6-10 |
| Study size | 10 | Explain how the study size was arrived at | 6-10 |
| Quantitative variables | 11 | Explain how quantitative variables were handled in the analyses. If applicable, describe which groupings were chosen and why | 10 |
| Statistical methods | 12 | (*a*) Describe all statistical methods, including those used to control for confounding | 10 |
|  |  | (*b*) Describe any methods used to examine subgroups and interactions | 10 |
|  |  | (*c*) Explain how missing data were addressed | 10 |
|  |  | (*d*) If applicable, explain how loss to follow-up was addressed | 10 |
|  |  | (*e*) Describe any sensitivity analyses | 10 |
| Results | | |  |
| Participants | 13* | (a) Report numbers of individuals at each stage of study—eg numbers potentially eligible, examined for eligibility, confirmed eligible, included in the study, completing follow-up, and analysed | 11 |
|  |  | (b) Give reasons for non-participation at each stage | N/A |
|  |  | (c) Consider use of a flow diagram | N/A |
| Descriptive data | 14* | (a) Give characteristics of study participants (eg demographic, clinical, social) and information on exposures and potential confounders | 11 |
|  |  | (b) Indicate number of participants with missing data for each variable of interest | 25 |
|  |  | (c) Summarise follow-up time (eg, average and total amount) | 11 |
| Outcome data | 15* | Report numbers of outcome events or summary measures over time | 11-13 |
| Main results | 16 | (*a*) Give unadjusted estimates and, if applicable, confounder-adjusted estimates and their precision (eg, 95% confidence interval). Make clear which confounders were adjusted for and why they were included | 11-13 |
|  |  | (*b*) Report category boundaries when continuous variables were categorized | 11-13 |
|  |  | (*c*) If relevant, consider translating estimates of relative risk into absolute risk for a meaningful time period | N/A |
| Other analyses | 17 | Report other analyses done—eg analyses of subgroups and interactions, and sensitivity analyses | 11-13 |
| Discussion | | |  |
| Key results | 18 | Summarise key results with reference to study objectives | 14 |
| Limitations | 19 | Discuss limitations of the study, taking into account sources of potential bias or imprecision. Discuss both direction and magnitude of any potential bias | 16-17 |
| Interpretation | 20 | Give a cautious overall interpretation of results considering objectives, limitations, multiplicity of analyses, results from similar studies, and other relevant evidence | 15-17 |
| Generalisability | 21 | Discuss the generalisability (external validity) of the study results | 15-17 |
| Other information | | |  |
| Funding | 22 | Give the source of funding and the role of the funders for the present study and, if applicable, for the original study on which the present article is based | 18 |

Supplementary table 1: ICD-10/Read2 Codes used to define diagnoses

| **Subcategory** | **ICD10** | **Read2** |
| --- | --- | --- |
| Asthma | J45, J46 | 173A., 173c., 173d., 1780., 1781., 1782., 1783., 1784., 1785., 1786., 1787., 1788., 1789., 178A., 178B., 1O2.., 388t., 388t0, 38DL., 38DT., 38DV., 661M1, 661N1, 663.., 663d., 663e., 663e0, 663e1, 663f., 663j., 663m., 663N., 663n., 663N0, 663N1, 663N2, 663O0, 663P., 663p., 663P0, 663P1, 663P2, 663q., 663r., 663s., 663t., 663U., 663u., 663V., 663v., 663V0, 663V1, 663V2, 663V3, 663w., 663x., 663y., 66Y5., 66Y9., 66YA., 66YC., 66YJ., 66YK., 66YP., 66Yp., 66YQ., 66Yq., 66YR., 66Yr., 66Ys., 66Yu., 679J0, 679J1, 679J2, 8791., 8794., 8795., 8796., 8797., 8798., 8B3j., 8CMA0, 8CR0., 8H2P., 9NNX., 9OJ1., 9OJA., H3120, H33.., H330., H3300, H3301, H330z, H331., H3310, H3311, H331z, H333., H334., H335., H33z., H33z0, H33z1, H33z2, H33zz, H35y6, H35y7, H47y0 |
| COPD | J41, J42, J43, J44 | 661M3, 661N3, 663K., 66YB., 66YB0, 66YB1, 66YB2, 66YD., 66Yd., 66Ye., 66Yf., 66Yg., 66Yh., 66YI., 66Yi., 66YL., 66YM., 66YS., 66YT., 66Yz2, 679V., 8CMW5, 8CR1., 8H2R., 9NgP., 9Oi.., 9Oi0., 9Oi1., 9Oi2., 9Oi3., 9Oi4., H3..., H30z., H31.., H310., H3100, H310z, H311., H311z, H312., H3120, H3121, H3122, H312z, H31y., H31yz, H31z., H32.., H320., H3200, H3201, H3203, H320z, H321., H322., H32y., H32yz, H32z., H36.., H37.., H38.., H39.., H3A.., H3B.., H3y.., H3y0., H3y1., H3z.., Hyu30, Hyu31 |
| Heart failure | I11, I13, I50 | 14A6., 14AM., 1O1.., 388D., 661M5, 662f., 662g., 662h., 662i., 662p., 662T., 662W., 679W1, 679X., 8B29., 8CeC., 8CL3., 8CMK., 8CMW8, 8H2S., 8HBE., 8HHz., 8Hk0., 9h1.., 9h11., 9h12., 9hH.., 9hH0., 9hH1., 9N2p., 9N6T., 9On.., 9On0., 9On1., 9On2., 9On3., 9On4., 9Or.., 9Or0., 9Or1., 9Or2., 9Or3., 9Or4., 9Or5., G1yz1, G2101, G2111, G21z1, G232., G234., G400., G41z., G5540, G58.., G580., G5800, G5801, G5802, G5803, G5804, G581., G5810, G582., G584., G58z., G5yy9, G5yyA, ZRad. |
| Hypertension | I10, I11, I12, I13, I15 | 14A2., 21261, 212K., 61462, 6624., 6627., 6628., 662b., 662c., 662d., 662F., 662G., 662O., 662r., 7Q01., 8B26., 8BL0., 8I3N., 9OI9., F4042, F4213, G2..., G20.., G200., G201., G202., G203., G20z., G21.., G210., G2100, G2101, G211., G2110, G2111, G21z., G21z0, G21z1, G21zz, G22.., G220., G221., G222., G22z., G23.., G230., G231., G232., G233., G234., G23z., G24.., G240., G2400, G240z, G241., G2410, G241z, G244., G24z., G24z0, G24z1, G24zz, G2y.., G2z.., G672., Gyu2., Gyu21, L122., L1220, L1221, L1223, L122z, L127., L127z, L128., L1280, L1282, TJC7., TJC7z, U60C5 |
| Myocardial infarction | I21, I22 | 14A3., 14A4., 14AH., 323.., 3233., 3234., 3235., 3236., 323Z., 889A., G30.., G300., G301., G3010, G3011, G301z, G302., G303., G304., G305., G306., G307., G3070, G3071, G308., G309., G30A., G30B., G30X., G30X0, G30y., G30y0, G30y1, G30y2, G30yz, G30z., G310., G31y1, G32.., G33z5, G35.., G350., G351., G353., G35X., G36.., G360., G361., G362., G363., G364., G365., G366., G38.., G380., G381., G384., G38z., G501., Gyu34 |
| Stroke | G46, I63, I64 | 14A7., 14AK., 1M4.., 661M7, 661N7, 662e., 662M., 662M1, 662M2, 7P242, 8HHM., 8IEC., 9h2.., 9h21., 9h22., Fyu56, G63.., G63y0, G63y1, G64.., G640., G6400, G641., G6410, G64z., G64z0, G64z1, G64z2, G64z3, G64z4, G66.., G663., G664., G665., G666., G667., G668., G683., G68X., G6W.., G6X.., Gyu63, Gyu64, Gyu6C, Gyu6G, L440., ZV125 |

Supplementary Table 2: BNF Codes used to define pharmacological management

| **Category** | **First 4 BNF code digits** |
| --- | --- |
| Beta blockers | 0204 |
| RAAS inhibitors | 0205 |
| Anticoagulants | 0208 |
| Antiplatelets | 0209 |
| Lipid lowering therapy | 0212 |

Supplementary Table 3: OPCS-4 Codes used to define revascularization procedures.

| **Category** | **OPCS4_Code** |
| --- | --- |
| Invasive angiography | K51, K63, K65 |
| Coronary artery bypass grafting | K40, K41, K42, K43, K44, K45, K46 |
| Percutaneous coronary intervention | K49, K50, K75 |

#### Supplementary Table 4 - Baseline characteristics of study population on first presentation per year

| **Characteristic** | **Overall**, N=164 572*^1^* | **2014**, N=12 026*^1^* | **2015**, N=12 077*^1^* | **2016**, N=13 105*^1^* | **2017**, N=14 414*^1^* | **2018**, N=15 388*^1^* | **2019**, N=15 795*^1^* | **2020**, N=15 076*^1^* | **2021**, N=17 335*^1^* | **2022**, N=16 480*^1^* | **2023**, N=16 490*^1^* | **2024**, N=16 386*^1^* |
| --- | --- | --- | --- | --- | --- | --- | --- | --- | --- | --- | --- | --- |
| **Age, yrs - mean (sd)** | 59 (18) | 61 (18) | 61 (18) | 61 (17) | 60 (18) | 60 (18) | 59 (18) | 59 (18) | 58 (18) | 59 (18) | 59 (18) | 58 (18) |
| **Sex, female** | 78 623 (48%) | 5 633 (47%) | 5 603 (46%) | 6 265 (48%) | 6 861 (48%) | 7 325 (48%) | 7 497 (47%) | 7 222 (48%) | 8 478 (49%) | 7 843 (48%) | 7 963 (48%) | 7 933 (48%) |
| **SIMD** |  |  |  |  |  |  |  |  |  |  |  |  |
| *Quintile 1 (Most deprived)* | 27 407 (17%) | 2 026 (17%) | 2 037 (17%) | 2 230 (17%) | 2 437 (17%) | 2 513 (16%) | 2 685 (17%) | 2 503 (17%) | 2 912 (17%) | 2 694 (16%) | 2 659 (16%) | 2 711 (17%) |
| *Quintile 2-4* | 98 484 (60%) | 7 111 (59%) | 7 226 (60%) | 7 770 (59%) | 8 531 (59%) | 9 251 (60%) | 9 399 (60%) | 9 138 (61%) | 10 463 (60%) | 9 863 (60%) | 9 875 (60%) | 9 857 (60%) |
| *Quintile 5 (Least deprived)* | 38 331 (23%) | 2 863 (24%) | 2 780 (23%) | 3 073 (24%) | 3 420 (24%) | 3 586 (23%) | 3 683 (23%) | 3 417 (23%) | 3 926 (23%) | 3 884 (24%) | 3 930 (24%) | 3 769 (23%) |
| *(Missing)* | 350 | 30 | 30 | 30 | 30 | 40 | 30 | 20 | 30 | 40 | 30 | 50 |
| **Ethnicity** |  |  |  |  |  |  |  |  |  |  |  |  |
| *Asian* | 5 368 (3·4%) | 287 (2·4%) | 314 (2·7%) | 342 (2·7%) | 423 (3·1%) | 473 (3·2%) | 483 (3·2%) | 468 (3·3%) | 632 (3·9%) | 620 (4·0%) | 653 (4·3%) | 673 (4·5%) |
| *Black* | 1 323 (0·8%) | 40 (0·3%) | 57 (0·5%) | 69 (0·5%) | 76 (0·5%) | 98 (0·7%) | 119 (0·8%) | 121 (0·8%) | 184 (1·1%) | 158 (1·0%) | 171 (1·1%) | 230 (1·5%) |
| *Other/Unknown* | 7 774 (5·0%) | 443 (3·8%) | 428 (3·6%) | 522 (4·1%) | 633 (4·6%) | 669 (4·5%) | 692 (4·6%) | 719 (5·0%) | 944 (5·8%) | 883 (5·7%) | 887 (5·8%) | 954 (6·3%) |
| *White* | 141 859 (91%) | 10 972 (93%) | 10 939 (93%) | 11 770 (93%) | 12 710 (92%) | 13 465 (92%) | 13 726 (91%) | 12 970 (91%) | 14 614 (89%) | 13 797 (89%) | 13 631 (89%) | 13 265 (88%) |
| *(Missing)* | 8 250 | 280 | 340 | 400 | 570 | 680 | 780 | 800 | 960 | 1 020 | 1 150 | 1 260 |
| **Body mass index** | 28 (24 – 32) | 27 (24 – 32) | 28 (24 – 32) | 28 (24 – 32) | 28 (24 – 32) | 28 (24 – 32) | 28 (24 – 32) | 28 (24 – 32) | 28 (24 – 32) | 28 (24 – 32) | 28 (24 – 32) | 27 (24 – 32) |
| **Smoking** |  |  |  |  |  |  |  |  |  |  |  |  |
| *Current smoker* | 34 832 (23%) | 2 687 (24%) | 2 675 (24%) | 2 874 (24%) | 3 164 (24%) | 3 285 (23%) | 3 429 (24%) | 3 094 (22%) | 3 659 (23%) | 3 313 (22%) | 3 344 (22%) | 3 308 (22%) |
| *Ex smoker* | 66 385 (44%) | 5 085 (46%) | 5 121 (46%) | 5 442 (45%) | 5 965 (45%) | 6 375 (45%) | 6 333 (44%) | 6 184 (45%) | 6 830 (43%) | 6 499 (43%) | 6 368 (42%) | 6 183 (41%) |
| *Non smoker* | 49 991 (33%) | 3 285 (30%) | 3 324 (30%) | 3 779 (31%) | 4 117 (31%) | 4 486 (32%) | 4 730 (33%) | 4 609 (33%) | 5 474 (34%) | 5 357 (35%) | 5 336 (35%) | 5 494 (37%) |
| **eGFR [mL/min/1.73m^2]** |  |  |  |  |  |  |  |  |  |  |  |  |
| *>60* | 134 350 (82%) | 9 503 (79%) | 9 688 (80%) | 10 638 (81%) | 11 799 (82%) | 12 627 (82%) | 12 913 (82%) | 12 341 (82%) | 14 438 (84%) | 13 441 (82%) | 13 540 (82%) | 13 422 (82%) |
| *30-60* | 25 356 (15%) | 2 106 (18%) | 1 988 (16%) | 2 062 (16%) | 2 163 (15%) | 2 324 (15%) | 2 418 (15%) | 2 316 (15%) | 2 396 (14%) | 2 538 (15%) | 2 513 (15%) | 2 532 (16%) |
| *<30* | 4 299 (2·6%) | 366 (3·1%) | 374 (3·1%) | 357 (2·7%) | 414 (2·9%) | 410 (2·7%) | 431 (2·7%) | 392 (2·6%) | 436 (2·5%) | 408 (2·5%) | 369 (2·2%) | 342 (2·1%) |
| *(Missing)* | 567 | 51 | 27 | 48 | 38 | 27 | 33 | 27 | 65 | 93 | 68 | 90 |
| **Past medical history** | | | | | | | | | | | | |
| *Hypertension* | 57 279 (35%) | 4 665 (39%) | 4 661 (39%) | 4 859 (37%) | 5 212 (36%) | 5 565 (36%) | 5 456 (35%) | 5 042 (33%) | 5 547 (32%) | 5 431 (33%) | 5 455 (33%) | 5 386 (33%) |
| *Stroke* | 10 754 (6·5%) | 931 (7·7%) | 922 (7·6%) | 963 (7·3%) | 950 (6·6%) | 1 080 (7·0%) | 1 036 (6·6%) | 987 (6·5%) | 1 043 (6·0%) | 978 (5·9%) | 1 009 (6·1%) | 855 (5·2%) |
| *COPD_Asthma* | 42 753 (26%) | 3 035 (25%) | 3 112 (26%) | 3 359 (26%) | 3 647 (25%) | 4 083 (27%) | 4 098 (26%) | 3 968 (26%) | 4 527 (26%) | 4 236 (26%) | 4 326 (26%) | 4 362 (27%) |
| **Previous revascularisation** | | | | | | | | | | | | |
| *PCI* | 16 859 (10%) | 1 328 (11%) | 1 334 (11%) | 1 430 (11%) | 1 527 (11%) | 1 659 (11%) | 1 674 (11%) | 1 557 (10%) | 1 641 (9·5%) | 1 576 (9·6%) | 1 600 (9·7%) | 1 533 (9·4%) |
| *CABG* | 3 754 (2·3%) | 360 (3·0%) | 351 (2·9%) | 359 (2·7%) | 366 (2·5%) | 376 (2·4%) | 365 (2·3%) | 331 (2·2%) | 361 (2·1%) | 308 (1·9%) | 300 (1·8%) | 277 (1·7%) |
| **Prescribing (12 months prior to attendance)** | | | | | | | | | | | | |
| *Antiplatelet* | 43 557 (26%) | 4 170 (35%) | 4 047 (34%) | 4 013 (31%) | 4 068 (28%) | 4 200 (27%) | 4 099 (26%) | 3 758 (25%) | 3 965 (23%) | 3 850 (23%) | 3 816 (23%) | 3 571 (22%) |
| *Anticoagulant* | 16 406 (10·0%) | 951 (7·9%) | 1 076 (8·9%) | 1 155 (8·8%) | 1 311 (9·1%) | 1 561 (10%) | 1 589 (10%) | 1 555 (10%) | 1 751 (10%) | 1 773 (11%) | 1 836 (11%) | 1 848 (11%) |
| *Lipid regulating drug* | 60 022 (36%) | 4 839 (40%) | 4 898 (41%) | 4 986 (38%) | 5 373 (37%) | 5 601 (36%) | 5 689 (36%) | 5 394 (36%) | 5 789 (33%) | 5 706 (35%) | 5 925 (36%) | 5 822 (36%) |
| *Beta blocker* | 41 014 (25%) | 3 323 (28%) | 3 381 (28%) | 3 411 (26%) | 3 722 (26%) | 3 886 (25%) | 3 830 (24%) | 3 679 (24%) | 3 959 (23%) | 3 985 (24%) | 3 985 (24%) | 3 853 (24%) |
| *Renin-angiotensin system drug* | 49 323 (30%) | 4 105 (34%) | 4 053 (34%) | 4 179 (32%) | 4 532 (31%) | 4 735 (31%) | 4 635 (29%) | 4 415 (29%) | 4 689 (27%) | 4 678 (28%) | 4 655 (28%) | 4 647 (28%) |
| *^1^*Mean (SD); n (%) | | | | | | | | | | | | |

#### Supplementary Table 5 – Cardiovascular outcomes in patients not diagnosed with myocardial infarction at first presentation each year

| **Characteristic** | **Overall**, N=109 457*^1^* | **2014**, N=11 104*^1^* | **2015**, N=9 558*^1^* | **2016**, N=9 551*^1^* | **2017**, N=10 156*^1^* | **2018**, N=10 251*^1^* | **2019**, N=10 195*^1^* | **2020**, N=9 341*^1^* | **2021**, N=10 649*^1^* | **2022**, N=9 708*^1^* | **2023**, N=9 639*^1^* | **2024**, N=9 305*^1^* |
| --- | --- | --- | --- | --- | --- | --- | --- | --- | --- | --- | --- | --- |
| *Admitted* | 42 597 (39%) | 6 163 (56%) | 4 753 (50%) | 3 241 (34%) | 3 462 (34%) | 3 429 (33%) | 3 821 (37%) | 3 891 (42%) | 4 037 (38%) | 3 159 (33%) | 3 478 (36%) | 3 163 (34%) |
| *ED (hours)* | 4 (3 – 5) | 4 (3 – 4) | 4 (3 – 4) | 4 (3 – 4) | 4 (3 – 4) | 4 (3 – 4) | 3 (3 – 4) | 3 (3 – 4) | 4 (3 – 5) | 6 (4 – 8) | 5 (4 – 7) | 6 (4 – 8) |
| *ED + inpatient (days)* | 0 (0 – 1) | 1 (0 – 2) | 0 (0 – 1) | 0 (0 – 1) | 0 (0 – 1) | 0 (0 – 1) | 0 (0 – 1) | 0 (0 – 1) | 0 (0 – 1) | 0 (0 – 1) | 0 (0 – 1) | 0 (0 – 1) |
| *Angiography* | 2 386 (2·2%) | 294 (2·6%) | 252 (2·6%) | 208 (2·2%) | 227 (2·2%) | 232 (2·3%) | 230 (2·3%) | 217 (2·3%) | 166 (1·6%) | 194 (2·0%) | 206 (2·1%) | 160 (1·7%) |
| **Revascularisation within 30 days from attendance** | | | | | | | | | | | | |
| *PCI* | 1 020 (0·9%) | 111 (1·0%) | 114 (1·2%) | 87 (0·9%) | 100 (1·0%) | 101 (1·0%) | 93 (0·9%) | 100 (1·1%) | 69 (0·6%) | 78 (0·8%) | 97 (1·0%) | 70 (0·8%) |
| *CABG* | 187 (0·2%) | 28 (0·3%) | 6 (<0·1%) | 17 (0·2%) | 15 (0·1%) | 13 (0·1%) | 19 (0·2%) | 14 (0·1%) | 13 (0·1%) | 31 (0·3%) | 17 (0·2%) | 14 (0·2%) |
| **Any reattendance from discharge** | | | | | | | | | | | | |
| *30 days* | 13 052 (12%) | 1 576 (14%) | 1 215 (13%) | 1 110 (12%) | 1 223 (12%) | 1 262 (12%) | 1 217 (12%) | 1 076 (12%) | 1 221 (11%) | 1 058 (11%) | 1 084 (11%) | 1 010 (11%) |
| *1 year* | 42 797 (39%) | 5 082 (46%) | 4 134 (43%) | 3 870 (41%) | 4 055 (40%) | 4 136 (40%) | 3 905 (38%) | 3 516 (38%) | 4 046 (38%) | 3 580 (37%) | 3 579 (37%) | 2 894 (31%) |
| **Reattendance with suspected ACS from discharge** | | | | | | | | | | | | |
| *30 days* | 4 254 (3·9%) | 598 (5·4%) | 400 (4·2%) | 337 (3·5%) | 423 (4·2%) | 385 (3·8%) | 384 (3·8%) | 350 (3·7%) | 416 (3·9%) | 315 (3·2%) | 334 (3·5%) | 312 (3·4%) |
| *1 year* | 13 068 (12%) | 1 937 (17%) | 1 361 (14%) | 1 201 (13%) | 1 263 (12%) | 1 225 (12%) | 1 124 (11%) | 1 143 (12%) | 1 215 (11%) | 1 018 (10%) | 993 (10%) | 588 (6·3%) |
| **Reattendance with MI from discharge** | | | | | | | | | | | | |
| *30 days* | 203 (0·2%) | 26 (0·2%) | 20 (0·2%) | 15 (0·2%) | 12 (0·1%) | 24 (0·2%) | 20 (0·2%) | 21 (0·2%) | 8 (<0·1%) | 22 (0·2%) | 21 (0·2%) | 14 (0·2%) |
| *1 year* | 667 (0·6%) | 121 (1·1%) | 76 (0·8%) | 60 (0·6%) | 56 (0·6%) | 69 (0·7%) | 71 (0·7%) | 50 (0·5%) | 43 (0·4%) | 40 (0·4%) | 54 (0·6%) | 27 (0·3%) |
| *within study period* | 2 192 (2·0%) | 506 (4·6%) | 373 (3·9%) | 277 (2·9%) | 269 (2·6%) | 212 (2·1%) | 177 (1·7%) | 123 (1·3%) | 92 (0·9%) | 74 (0·8%) | 62 (0·6%) | 27 (0·3%) |
| **Death (all-cause) from attendance** | | | | | | | | | | | | |
| *inpatient* | 1 820 (1·7%) | 276 (2·5%) | 188 (2·0%) | 143 (1·5%) | 163 (1·6%) | 174 (1·7%) | 137 (1·3%) | 136 (1·5%) | 170 (1·6%) | 165 (1·7%) | 140 (1·5%) | 128 (1·4%) |
| *30 days* | 2 354 (2·2%) | 341 (3·1%) | 241 (2·5%) | 200 (2·1%) | 216 (2·1%) | 219 (2·1%) | 181 (1·8%) | 189 (2·0%) | 217 (2·0%) | 208 (2·1%) | 184 (1·9%) | 158 (1·7%) |
| *1 year* | 5 284 (4·8%) | 809 (7·3%) | 587 (6·1%) | 495 (5·2%) | 501 (4·9%) | 461 (4·5%) | 441 (4·3%) | 412 (4·4%) | 472 (4·4%) | 414 (4·3%) | 399 (4·1%) | 293 (3·1%) |
| **Death (cardiovascular) from attendance** | | | | | | | | | | | | |
| *inpatient* | 1 068 (1·0%) | 146 (1·3%) | 96 (1·0%) | 80 (0·8%) | 93 (0·9%) | 113 (1·1%) | 92 (0·9%) | 81 (0·9%) | 101 (0·9%) | 102 (1·1%) | 83 (0·9%) | 81 (0·9%) |
| *30 days* | 1 359 (1·2%) | 178 (1·6%) | 116 (1·2%) | 108 (1·1%) | 132 (1·3%) | 136 (1·3%) | 118 (1·2%) | 110 (1·2%) | 131 (1·2%) | 125 (1·3%) | 106 (1·1%) | 99 (1·1%) |
| *1 year* | 2 832 (2·6%) | 398 (3·6%) | 267 (2·8%) | 246 (2·6%) | 281 (2·8%) | 263 (2·6%) | 261 (2·6%) | 228 (2·4%) | 270 (2·5%) | 225 (2·3%) | 220 (2·3%) | 173 (1·9%) |
| **Death (cardiac) from attendance** | | | | | | | | | | | | |
| *inpatient* | 720 (0·7%) | 104 (0·9%) | 61 (0·6%) | 54 (0·6%) | 71 (0·7%) | 72 (0·7%) | 58 (0·6%) | 62 (0·7%) | 63 (0·6%) | 67 (0·7%) | 55 (0·6%) | 53 (0·6%) |
| *30 days* | 936 (0·9%) | 129 (1·2%) | 76 (0·8%) | 76 (0·8%) | 100 (1·0%) | 89 (0·9%) | 75 (0·7%) | 84 (0·9%) | 83 (0·8%) | 85 (0·9%) | 74 (0·8%) | 65 (0·7%) |
| *1 year* | 1 967 (1·8%) | 294 (2·6%) | 180 (1·9%) | 178 (1·9%) | 208 (2·0%) | 176 (1·7%) | 171 (1·7%) | 171 (1·8%) | 178 (1·7%) | 152 (1·6%) | 147 (1·5%) | 112 (1·2%) |
| *^1^*n (%); Median (IQR) | | | | | | | | | | | | |

#### Supplementary Table 6 – Cardiovascular outcomes in patients diagnosed with myocardial infarction at first presentation each year

| **Characteristic** | **Overall**, N=7 685*^1^* | **2014**, N=922*^1^* | **2015**, N=808*^1^* | **2016**, N=780*^1^* | **2017**, N=787*^1^* | **2018**, N=770*^1^* | **2019**, N=681*^1^* | **2020**, N=667*^1^* | **2021**, N=666*^1^* | **2022**, N=570*^1^* | **2023**, N=509*^1^* | **2024**, N=525*^1^* |
| --- | --- | --- | --- | --- | --- | --- | --- | --- | --- | --- | --- | --- |
| *Admitted* | 7 685 (100%) | 922 (100%) | 808 (100%) | 780 (100%) | 787 (100%) | 770 (100%) | 681 (100%) | 667 (100%) | 666 (100%) | 570 (100%) | 509 (100%) | 525 (100%) |
| *ED (hours)* | 4 (3 – 5) | 3 (3 – 4) | 3 (3 – 4) | 4 (3 – 4) | 3 (3 – 4) | 3 (2 – 5) | 4 (3 – 5) | 3 (2 – 4) | 4 (3 – 6) | 7 (4 – 12) | 6 (3 – 10) | 7 (3 – 12) |
| *ED + inpatient (days)* | 3 (2 – 5) | 3 (2 – 5) | 3 (2 – 6) | 3 (2 – 6) | 3 (2 – 5) | 3 (2 – 5) | 3 (2 – 5) | 3 (2 – 5) | 3 (2 – 5) | 3 (2 – 6) | 3 (2 – 5) | 3 (2 – 5) |
| *Angiography* | 5 546 (72%) | 631 (68%) | 559 (69%) | 553 (71%) | 553 (70%) | 559 (73%) | 511 (75%) | 490 (73%) | 504 (76%) | 402 (71%) | 379 (74%) | 405 (77%) |
| **Revascularisation within 30 days from attendance** | | | | | | | | | | | | |
| *PCI* | 4 227 (55%) | 471 (51%) | 429 (53%) | 417 (53%) | 451 (57%) | 437 (57%) | 380 (56%) | 371 (56%) | 372 (56%) | 304 (53%) | 285 (56%) | 310 (59%) |
| *CABG* | 285 (3·7%) | 33 (3·6%) | 25 (3·1%) | 27 (3·5%) | 25 (3·2%) | 28 (3·6%) | 23 (3·4%) | 31 (4·6%) | 31 (4·7%) | 23 (4·0%) | 16 (3·1%) | 23 (4·4%) |
| **Any reattendance from discharge** | | | | | | | | | | | | |
| *30 days* | 1 327 (17%) | 177 (19%) | 158 (20%) | 154 (20%) | 125 (16%) | 131 (17%) | 113 (17%) | 126 (19%) | 96 (14%) | 83 (15%) | 84 (17%) | 80 (15%) |
| *1 year* | 3 321 (43%) | 426 (46%) | 384 (48%) | 372 (48%) | 346 (44%) | 338 (44%) | 297 (44%) | 295 (44%) | 257 (39%) | 214 (38%) | 216 (42%) | 176 (34%) |
| *within study period* | 711 (9·3%) | 141 (15%) | 106 (13%) | 95 (12%) | 91 (12%) | 77 (10%) | 57 (8·4%) | 47 (7·0%) | 48 (7·2%) | 27 (4·7%) | 16 (3·1%) | 6 (1·1%) |
| **Reattendance with suspected ACS from discharge** | | | | | | | | | | | | |
| *30 days* | 912 (12%) | 117 (13%) | 110 (14%) | 111 (14%) | 86 (11%) | 89 (12%) | 69 (10%) | 95 (14%) | 72 (11%) | 58 (10%) | 51 (10%) | 54 (10%) |
| *1 year* | 2 049 (27%) | 259 (28%) | 262 (32%) | 236 (30%) | 215 (27%) | 202 (26%) | 176 (26%) | 199 (30%) | 154 (23%) | 141 (25%) | 121 (24%) | 84 (16%) |
| **Reattendance with MI from discharge** | | | | | | | | | | | | |
| *30 days* |  | 18 (2·0%) | 18 (2·2%) | 19 (2·4%) | 9 (1·1%) | 12 (1·6%) | 9 (1·3%) | 13 (1·9%) | 15 (2·3%) | 6 (1·1%) | 9 (1·8%) | <5 (0·6%) |
| *1 year* | 315 (4·1%) | 49 (5·3%) | 37 (4·6%) | 36 (4·6%) | 38 (4·8%) | 31 (4·0%) | 27 (4·0%) | 29 (4·3%) | 30 (4·5%) | 18 (3·2%) | 14 (2·8%) | 6 (1·1%) |
| **Death (all-cause) from attendance** | | | | | | | | | | | | |
| *inpatient* | 499 (6·5%) | 77 (8·4%) | 58 (7·2%) | 47 (6·0%) | 61 (7·8%) | 51 (6·6%) | 31 (4·6%) | 38 (5·7%) | 46 (6·9%) | 29 (5·1%) | 34 (6·7%) | 27 (5·1%) |
| *30 days* | 621 (8·1%) | 98 (11%) | 71 (8·8%) | 56 (7·2%) | 73 (9·3%) | 59 (7·7%) | 46 (6·8%) | 47 (7·0%) | 58 (8·7%) | 35 (6·1%) | 45 (8·8%) | 33 (6·3%) |
| *1 year* | 942 (12%) | 149 (16%) | 114 (14%) | 83 (11%) | 112 (14%) | 86 (11%) | 74 (11%) | 75 (11%) | 81 (12%) | 56 (9·8%) | 63 (12%) | 49 (9·3%) |
| **Death (cardiovascular) from attendance** | | | | | | | | | | | | |
| *inpatient* | 447 (5·8%) | 66 (7·2%) | 49 (6·1%) | 44 (5·6%) | 56 (7·1%) | 48 (6·2%) | 27 (4·0%) | 31 (4·6%) | 40 (6·0%) | 28 (4·9%) | 31 (6·1%) | 27 (5·1%) |
| *30 days* | 548 (7·1%) | 86 (9·3%) | 59 (7·3%) | 51 (6·5%) | 65 (8·3%) | 55 (7·1%) | 40 (5·9%) | 37 (5·5%) | 49 (7·4%) | 32 (5·6%) | 41 (8·1%) | 33 (6·3%) |
| *1 year* | 753 (9·8%) | 115 (12%) | 84 (10%) | 70 (9·0%) | 92 (12%) | 67 (8·7%) | 60 (8·8%) | 56 (8·4%) | 67 (10%) | 45 (7·9%) | 52 (10%) | 45 (8·6%) |
| **Death (cardiac) from attendance** | | | | | | | | | | | | |
| *inpatient* | 431 (5·6%) | 63 (6·8%) | 47 (5·8%) | 42 (5·4%) | 53 (6·7%) | 47 (6·1%) | 25 (3·7%) | 30 (4·5%) | 38 (5·7%) | 28 (4·9%) | 31 (6·1%) | 27 (5·1%) |
| *30 days* | 526 (6·8%) | 81 (8·8%) | 56 (6·9%) | 49 (6·3%) | 61 (7·8%) | 54 (7·0%) | 38 (5·6%) | 35 (5·2%) | 47 (7·1%) | 32 (5·6%) | 41 (8·1%) | 32 (6·1%) |
| *1 year* | 690 (9·0%) | 103 (11%) | 81 (10%) | 63 (8·1%) | 80 (10%) | 62 (8·1%) | 52 (7·6%) | 53 (7·9%) | 58 (8·7%) | 44 (7·7%) | 50 (9·8%) | 44 (8·4%) |
| *^1^*n (%); Median (IQR) | | | | | | | | | | | | |

Supplementary table 7 – Medication use within 12 months after myocardial infarction, stratified by treating specialty

| **Characteristic** | **Cardiology** N = 8 566*^1^* | **Non-Cardiology** N = 1 836*^1^* | **p-value***^2^* |
| --- | --- | --- | --- |
| Lipid regulating drug | 8 095 (95%) | 1 256 (68%) | <0·0001 |
| DAPT | 7 706 (90%) | 1 186 (65%) | <0·0001 |
| beta-blocker | 6 258 (73%) | 1 080 (59%) | <0·0001 |
| Renin-Angiotensin system drug | 6 882 (80%) | 954 (52%) | <0·0001 |
| *^1^*n (%) | | | |
| *^2^*Pearson's Chi-squared test | | | |

### Supplementary Figure 1 – Presenting symptom recorded by clinical care team upon requesting measurement of cardiac troponin.

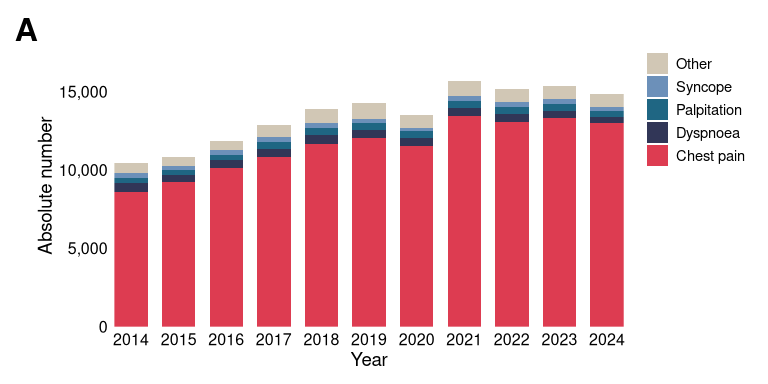

### Supplementary Figure 2 – Annual numbers of patients with myocardial infarction, stratified according to clinical diagnosis of STEMI or NSTEMI. Light blue bars indicates those patients under a non-cardiology treating specialty.

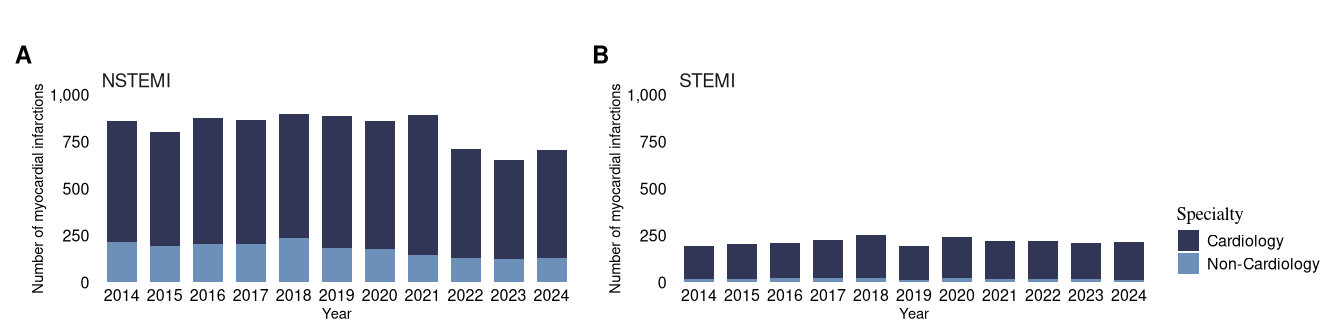
